# Advanced lung cancer inflammation index as a predictor of all-cause and cardiovascular mortality among American hypertensive patients

**DOI:** 10.1101/2024.10.22.24315955

**Authors:** Nan Hu, Yin Feng, Liqun He

**Affiliations:** Traditional Chinese and Western Medicine Hospital of Wuhan, Tongji Medical College, Huazhong University of Science and Technology. Wuhan, China

## Abstract

**BACKGROUND:** The predictive value of advanced lung cancer inflammation index (ALI) in all-cause mortality and cardiovascular mortality among hypertensive patients has not been thoroughly investigated.

**METHOD AND RESULTS:** A total of 2805 hypertensive patients from the National Health and Nutrition Examination Surveys from 2005-2006 to 2017-2018 were ultimately included in the analysis. Over the study period, participants were followed up for an average of 57.6 months, during which 311 deaths occurred, with 89 deaths attributed to cardiovascular causes. Survey-weighted restricted cubic spline (RCS) analysis revealed a nonlinear negative correlation between ALI and both all-cause and cardiovascular mortality among hypertensive patients. Survey-weighted Cox proportional hazards models revealed that in the highest ALI quartile experienced a 53.8% reduction in all-cause mortality and an 83.5% reduction in cardiovascular mortality compared with those in the lowest quartile. Survey-weighted Kaplan-Meier survival analysis also revealed that the all-cause mortality and cardiovascular mortality of hypertensive patients in the high ALI group were significantly lower than those in the low ALI group. Time-dependent ROC curves were used to assess the accuracy of survival outcomes and the area under the curve (AUC) values for the ability of ALI to predict all-cause mortality at 1, 3, and 5 years among hypertensive patients were 0.772, 0.672, and 0.634, respectively, whereas the AUC values for predicting cardiovascular mortality were 0.735, 0.760, and 0.723, respectively.

**CONCLUSIONS:** ALI can be a valuable and effective tool for identifying high-risk hypertensive patients and guiding targeted interventions.

## INTRODUCTION

Hypertension is a common chronic disease and is one of the main causes of death ^1^. In recent years, the incidence of hypertension has increased ^2^, and hypertension is widely recognized as a major risk factor for cardiovascular disease, stroke, and chronic kidney disease ^3-5^. Hypertension affects more than 1 billion people worldwide and directly causes nearly 10 million deaths ^6^. Reducing mortality and increasing life span are the primary goals of global public health ^7^. Therefore, a novel prognostic biomarker that can identify patients with high mortality at an early stage is needed.

The advanced lung cancer inflammation index (ALI) is calculated by multiplying the body mass index (BMI) by the serum ALB level and dividing it by the Neutrophil to Lymphocyte ratio (NLR) ^8^, and the NLR is determined by the ratio of neutrophils to lymphocytes ^9^. Previous studies have shown that ALI is associated with the occurrence and prognosis of multiple system disorders, including cardiovascular disease, stroke, diabetes, lung cancer, gastrointestinal cancer and rheumatoid arthritis ^10-15^. However, the associations between ALI and the risk of both all-cause mortality and cardiovascular mortality among hypertensive patients have not been fully elucidated.

In this analysis, we conducted a study across seven cycles of the National Health and Nutrition Examination Survey (NHANES), encompassing a vast population, with the aim of exploring the link between ALI and both all-cause mortality and cardiovascular mortality in hypertensive patients. This endeavor seeks to offer fresh perspectives on their treatment and prognosis, ultimately enhancing their quality of life and survival rates.

## METHODS

### Study population and design

The NHANES database, fully named the National Health and Nutrition Examination Survey, is a large-scale cross-sectional research project administered by the National Center for Health Statistics (NCHS) in the United States. This database aims to evaluate the health and nutritional status of adults and children in the United States, collecting comprehensive data through interviews and physical examinations, covering demographics, diet, healthcare, physiological measurements, laboratory tests, and mortality ^16^. More information about the NHANES database can be readily obtained from the official NHANES website (http://www.cdc.gov/nchs/nhanes/index.htm). Furthermore, all procedures adhered to the principles outlined in the Declaration of Helsinki, and NHANES has secured personal informed and written consent from every participant, ensuring their awareness and agreement in the study ^17^.

The following populations were screened out in this study: (1) younger than 18 years of age (n=28047); (2) missing data on hypertension (n=27800); (3) lacking information on survival status (n=27); (4) missing data on ALI measurements (n=1769); (5) lacking relevant information on any of the covariates(n=9620); and (6) outliers of ALI (n=123). A total of 2805 participants were ultimately included in the analysis, sourced from the 2005-2006 to 2017-2018 NHANES datasets (Supplemental Material: Figure S1).

### Assessment of hypertension

After a 5-minute break, participants’ blood pressure was measured three times consecutively and recorded as an average, and hypertension was defined as a blood pressure ≥ 140/90 mmHg ^18^. In this study, with the questionnaire “Have you ever been told by a doctor or other health professional that you have hypertension, also called high blood pressure?”, participants were classified as hypertensive if they answered “yes”.

### Measurement of ALI

The advanced lung cancer inflammation index (ALI) was measured via the equation “ALI = BMI × Alb/NLR”. BMI was measured by weight/height in kg/m². The NLR is calculated from the absolute neutrophil count/absolute lymphocyte count ^19^.

### Ascertainment of mortality

The main focus of this study was to assess all-cause mortality and cardiovascular mortality among hypertensive patients. To determine mortality among the follow-up population, we used the NHANES public-use mortality dataset, which has been linked to the National Death Index (NDI) by the National Center for Health Statistics (NCHS) and is current up to December 31, 2019. We employed the ICD-10 to classify deaths by heart disease (054-064), chronic lower respiratory diseases (082-086), and all other causes (010) ^20^.

### Covariates

Covariates such as age, sex, education, ethnicity, and the family poverty-to-income ratio can be obtained from “Demographics”. BMI was obtained from “Examination”, and we divided it into the categorization on the basis of the square of height in meters as follows: normal weight (less than 25 kg/m²), overweight (ranging from 25 to 30 kg/m²), and obese (30 kg/m² or above). ^21^. Triglyceride (TG), total cholesterol (TC), high-density lipoprotein (HDL), low-density lipoprotein, glycosylated haemoglobin (HbA1c), ALB, urinary creatinine (UCR) and the NLR. All data were obtained from the “laboratory”. In the “Questionnaire”, for smoking, when asked, “Have you smoked at least 100 cigarettes in your life?”, the participants answered “yes” or “no”. For diabetes, when asked, “Has a doctor or health professional ever told you diabetes or sugar diabetes except during pregnancy, have you diabetes or sugar diabetes?”, the participants also answered “yes” or “no”.

### Statistical analysis

Given the intricate multistage hierarchical probabilistic survey design employed by the NHANES, the statistical analysis incorporates a blend of sample weighting, clustering adjustments, and stratification to ensure the validity and representativeness of the study results ^22^. The Kruskal-Wallis test was used to compare continuous variables, whereas the chi-square test was used to compare categorical variables. In this study, ALI was categorized by machine learning methods: equal frequency binning ^23^.

To explore potential nonlinear associations between ALI and both all-cause mortality and cardiovascular mortality, particularly among hypertensive patients, we employed survey-weighted RCS analysis. On the basis of the minimum Akaike information criterion (AIC), the optimal nodes of all-cause mortality (3) and cardiovascular mortality (3) are determined ^24^.

Survey-weighted Cox proportional hazards models were used to evaluate the independent relationships of ALI with both all-cause mortality and cardiovascular mortality among hypertensive patients. To evaluate the independent predictive value of ALI, we constructed three models to present the findings. Model 1 was not adjusted. Model 2 was adjusted for age, sex, race, education and the family poverty income ratio. Model 3 was adjusted for age, sex, race, education, the family poverty income ratio, TG, TC, HDL, LDL, HbA1c, UCR, smoking status and diabetes status.

The optimal cut-off method involves segregating the continuous survival time data into two distinct groups to maximize the divergence between these groups. With this method, the optimal cut-off point was identified, and ALI was categorized into a high-level group and a low-level group ^25^. Survey-weighted Kaplan-Meier survival analysis was then employed to assess the survival probabilities of hypertensive patients and distinguish them on the basis of their ALI levels. For cardiovascular mortality, the Fine and Gray methods were used to analyze competing risk regression and cumulative incidence curves. ^26^.

Survey-weighted time-dependent ROC curves were used to assess the accuracy of survival outcomes predicted by ALI at 1 year, 3 years, and 5 years among hypertensive patients. Subgroup analyzes were performed according to age, sex, education, ethnicity, diabetes status, and smoking status. Notably, previous studies have shown that diabetes is associated with hypertension, all-cause mortality and cardiovascular mortality ^27-28^.

Statistical analyzes were conducted using R and DecisionLinnc Software. 2-sided p-value of less than 0.05 was considered statistically significant.

## RESULTS

### Participant characteristics

Table 1 presents the baseline characteristics of the cohort study participants (n = 2805) stratified by ALI quartile. The equal frequency binning divides ALI into four equal parts according to the number of participants: Q1 (4.14,44.02), Q2 (44.03,62.00), Q3 (62.02,84.04) and Q4 (84.08,151.61). Compared with the lowest quartile (Q1), ALI values were greater for the following participants: Seniors over the age of 65, non-Hispanic white, middle-income and obese patients. Additionally, with respect to the biochemical indices, TG, TC, LDL, ALB and UCR increased significantly with increasing quartile grade, whereas HDL levels tended to decrease (Table 1).

**Table 1.**
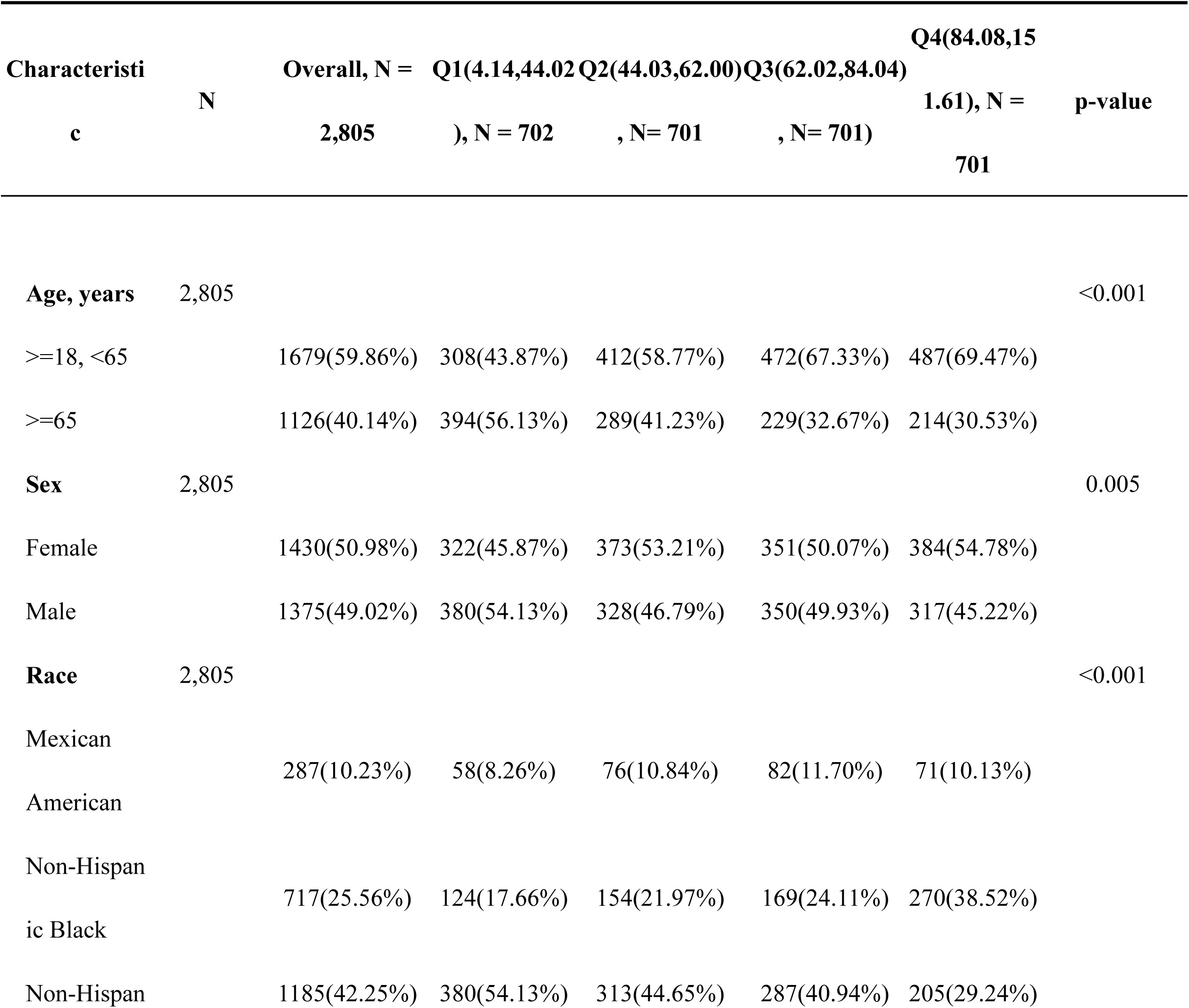
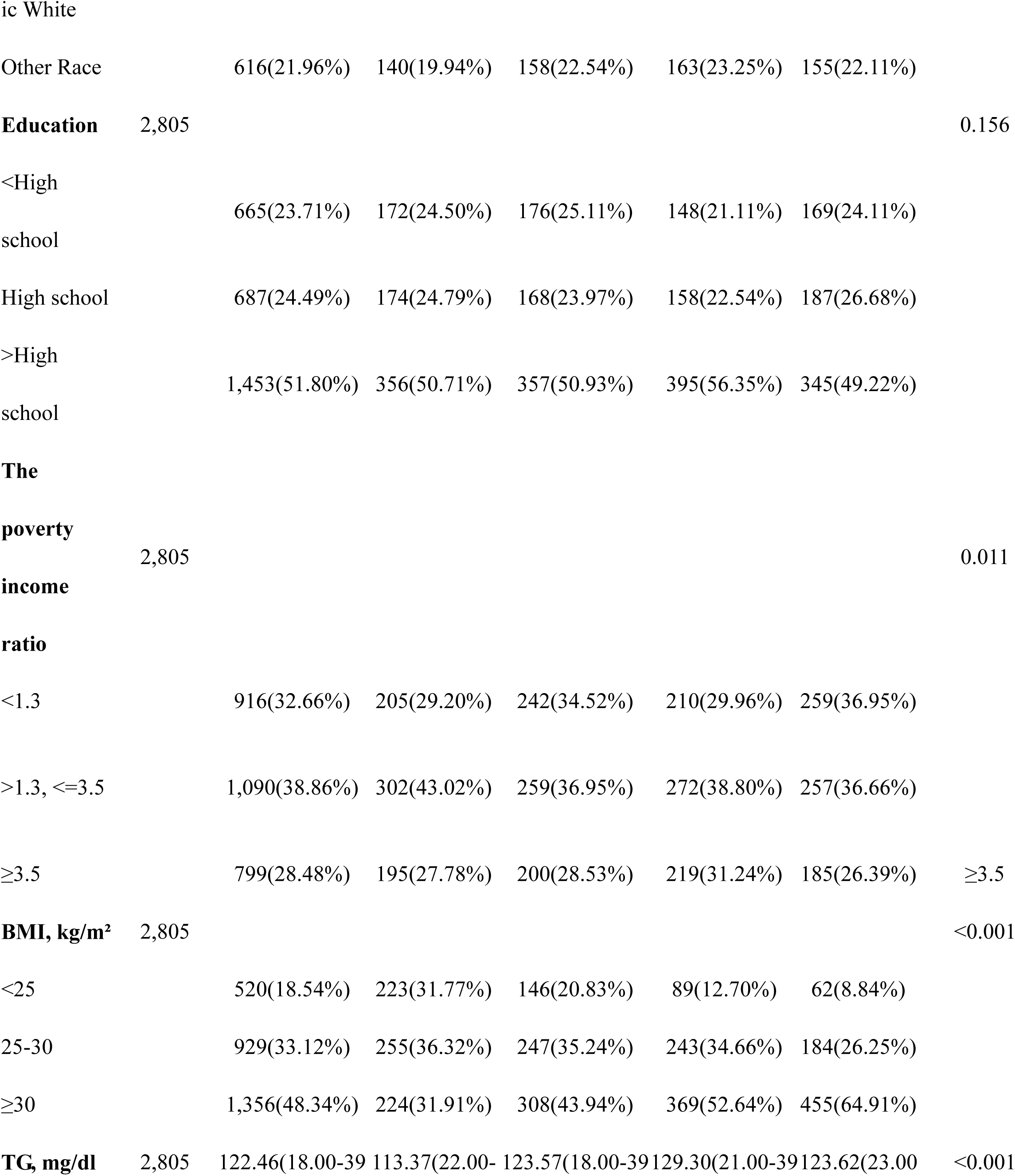
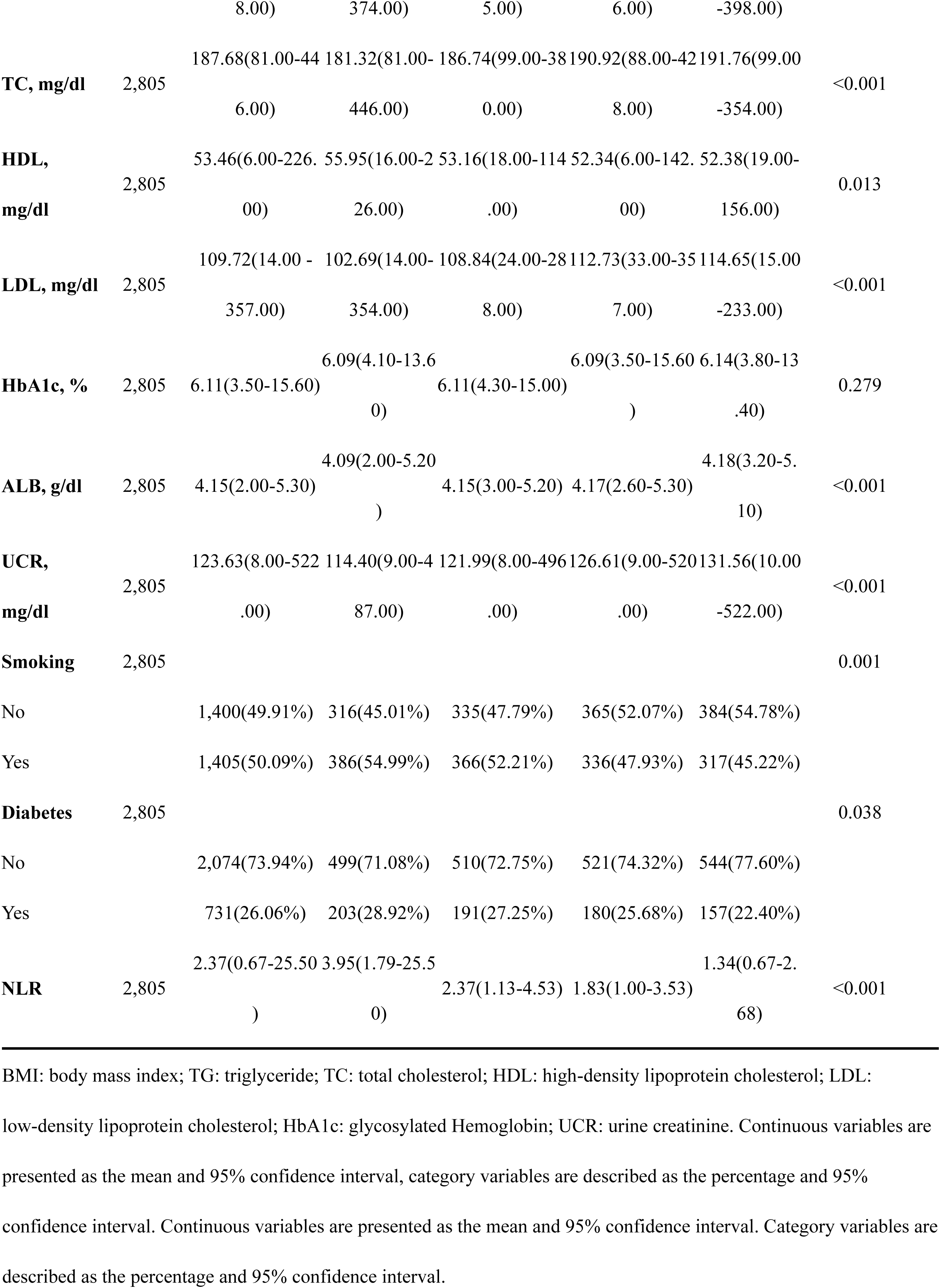
Baseline characteristics of 2805 hypertensive patients by quartiles of ALI index.

### Associations between ALI and all-cause mortality and cardiovascular mortality among hypertensive patients

In this study, during the average follow-up period of 57.6 (1,109) months, the total number of deaths was 311(11.09%), of which 89 (3.17%) died from cardiovascular causes. The RCS analysis revealed a nonlinear negative association between ALI and both all-cause mortality and cardiovascular mortality among hypertensive patients (p<0.001) (Figure 1); for details see the attached table (Supplemental Material: Table S1). Table 2 presents the independent associations between ALI and the risk of both all-cause mortality and cardiovascular death according to different Cox regression models. In Model 3, the hazard ratios (HRs), 95% confidence intervals (CIs) and p values for quartiles (Q1, Q2, Q3, and Q4) of all-cause mortality were 1.00(reference), 0.534(95%CI, 0.392-0.727; p<0.001), 0.488(95%CI, 0.346-0.690; p<0.001), and 0.462 (95%CI, 0.279-0.766; p=0.003), respectively. The cardiovascular mortality rates were 1.00(reference), 0.552(95%CI, 0.256-1.191; p=0.130), 0.327(95%CI, 0.147-0.725; p=0.006), and 0.165(95%CI, 0.056-0.482; p=0.001). After ALI reached the highest quartile, hypertensive patients experienced a 53.8% reduction in all-cause mortality risk and an 83.5% decrease in cardiovascular mortality risk (Table 2). Additionally, Cox regression analysis revealed a notable decline in mortality among those in the higher ALI quartiles (Q2, Q3, and Q4) compared with the lowest quartile (Q1), indicating a significant association between higher ALI and lower all-cause mortality. Moreover, this decreasing trend was more pronounced in the association of ALI with cardiovascular mortality among hypertensive patients.

**Figure 1.**
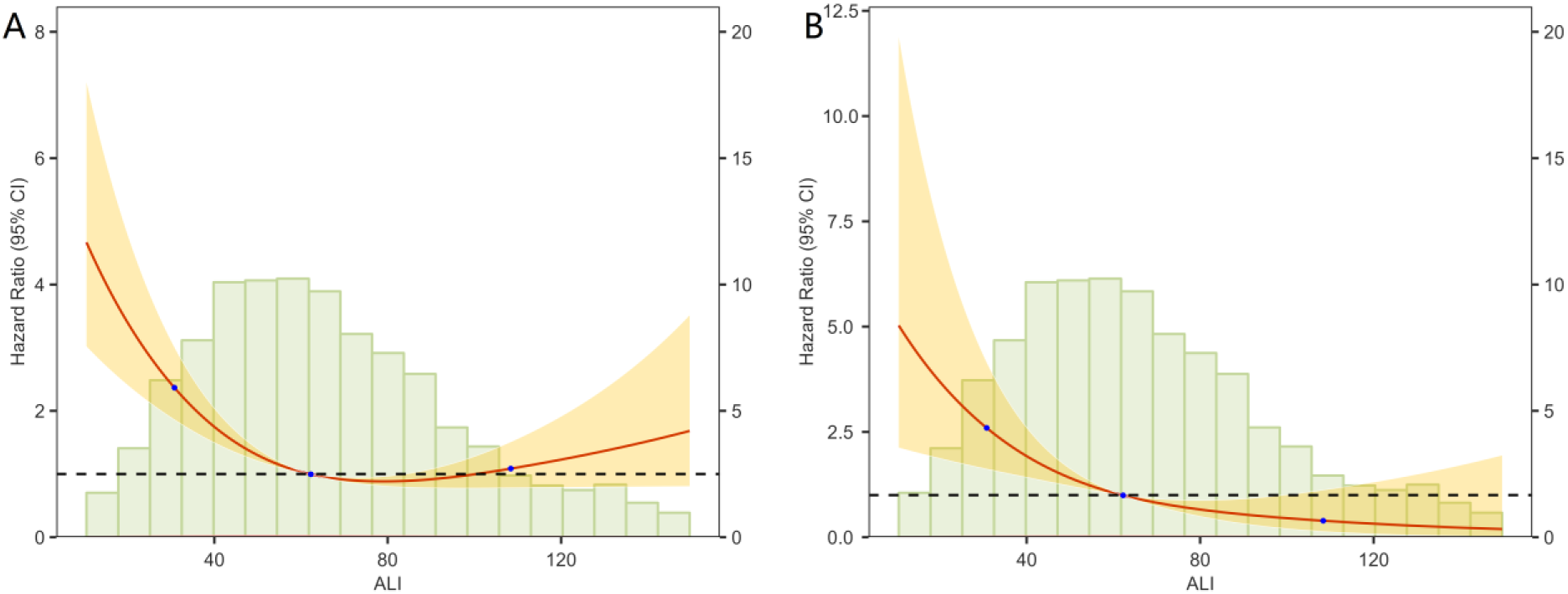
The association of Advanced lung cancer inflammation index (ALI) with all-cause mortality and cardiovascular mortality among 2805 hypertensive patients visualized by restricted cubic spline, weighted.

**Table 2.** HRs (95% CIs) for the mortality of 2805 hypertensive patients according to ALI quartiles.

| Characteristic | Model1 |  | Model2 |  | Model3 |  |
| --- | --- | --- | --- | --- | --- | --- |
|  | HR (95%CI) | P-value | HR (95%CI) | Pvalue | HR (95%CI) | P-value |
| **Overall** |  |  |  |  |  |  |
| **mortality** |  |  |  |  |  |  |
| Q1 | Reference |  | Reference |  | Reference |  |
| Q2 | 0.413(0.297,0.576) | <0.001 | 0.504(0.364,0.698) | <0.001 | 0.534(0.392,0.727) | <0.001 |
| Q3 | 0.325(0.230,0.459) | <0.001 | 0.440(0.304,0.637) | <0.001 | 0.488(0.346,0.690) | <0.001 |
| Q4 | 0.302(0.180,0.507) | <0.001 | 0.400(0.238,0.672) | 0.001 | 0.462(0.279,0.766) | 0.003 |
| **Cardiovascular** |  |  |  |  |  |  |
| **mortality** |  |  |  |  |  |  |

| Q1 | Reference |  | Reference |  | Reference |  |
| --- | --- | --- | --- | --- | --- | --- |
| Q2 | 0.454(0.221,0.931) | 0.031 | 0.587(0.279,1.234) | 0.160 | 0.552(0.256,1.191) | 0.130 |
| Q3 | 0.238(0.109,0.523) | <0.001 | 0.351(0.155,0.795) | 0.012 | 0.327(0.147,0.725) | 0.006 |
| Q4 | 0.120(0.042,0.344) | <0.001 | 0.166(0.057,0.486) | 0.001 | 0.165(0.056,0.482) | 0.001 |

Model1 : Unadjusted.
Model2 : Age、 Sex、 Education、 Race、 The poverty income ratio.
Model3 : Age、 Sex、 Education、 Race、 The poverty income ratio、 TG、 TC、 HDL、 LDL、 HbA1c、 UCR Smoking status and Diabetes status.
Weighted.

The Kaplan–Meier survival curve analysis also revealed that the all-cause mortality and cardiovascular mortality of hypertensive patients in the high ALI group were significantly lower than those in the low ALI group (Figure 2). The competing risk model presented the cumulative incidence function (CIF) of cardiovascular mortality and other-cause mortality at different time points. For both cardiovascular mortality and other-cause mortality, the incidence of mortality in the quartile gradually decreased (Q1>Q2>Q3>Q4) (Supplemental Material: Figure S2).

**Figure 2.**
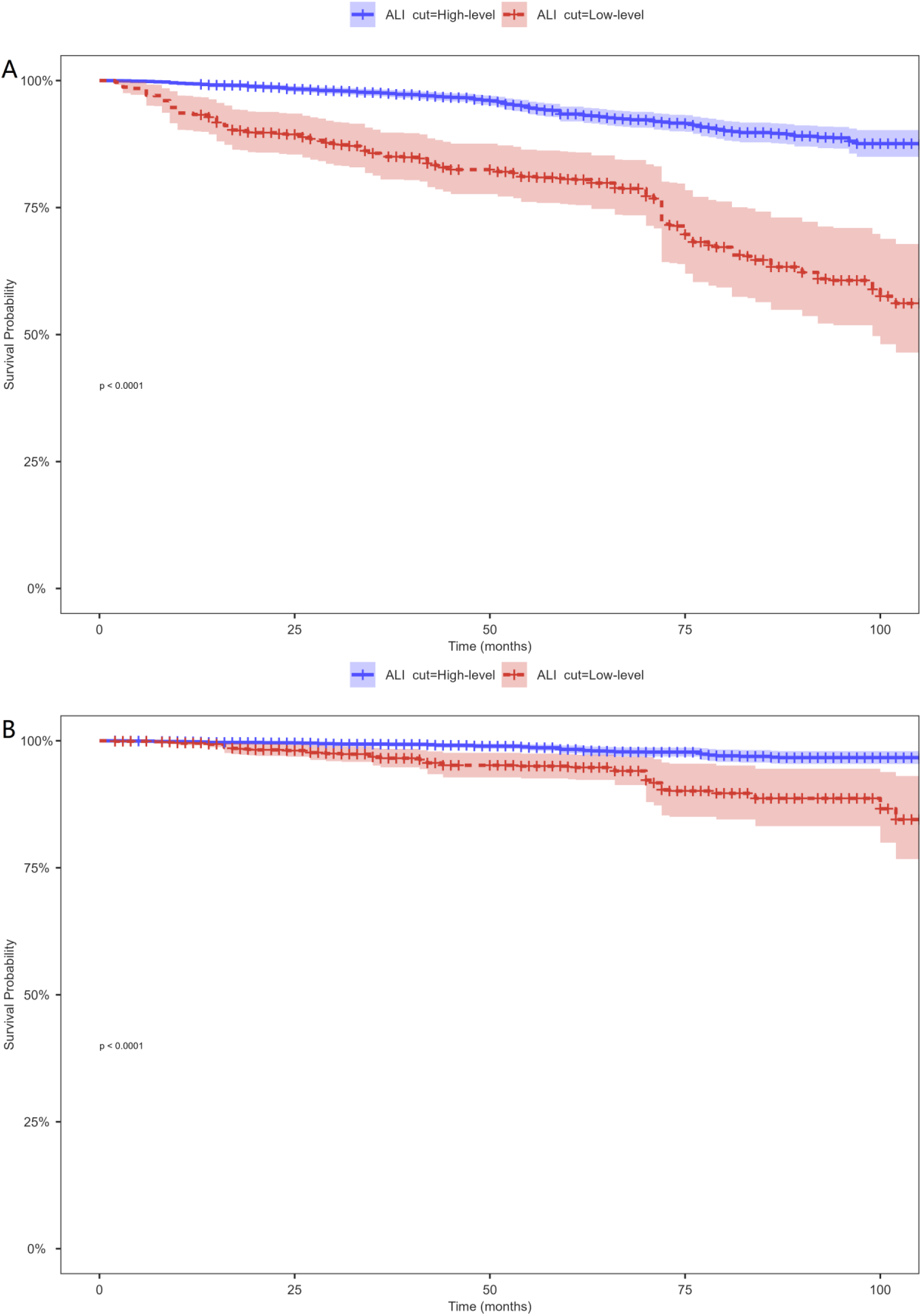
The survival rate with high-level (> 32.93) and low-level (≤ 32.93) Advanced lung cancer inflammation index (ALI) values among 2805 hypertensive patients, weighted.

### The ability of ALI to predict all-cause and cardiovascular death among hypertensive patients

Surveyed-weighted ROC curve analysis revealed that the areas under ALI curve (AUCs) for all-cause mortality at 1, 3, and 5 years were 0.772, 0.672 and 0.634, respectively, and those for cardiovascular mortality were 0.735, 0.760 and 0.723, respectively (Figure 3). These findings indicate that ALI is similar and capable of making effective forecasts for hypertensive patients across various time periods. Additionally, we analyzed the predictive power of the NLR for all-cause mortality and cardiovascular mortality among hypertensive patients. The results showed that the NLR is less effective in predicting mortality than ALI (Supplemental Material: Figure S3).

**Figure 3.**
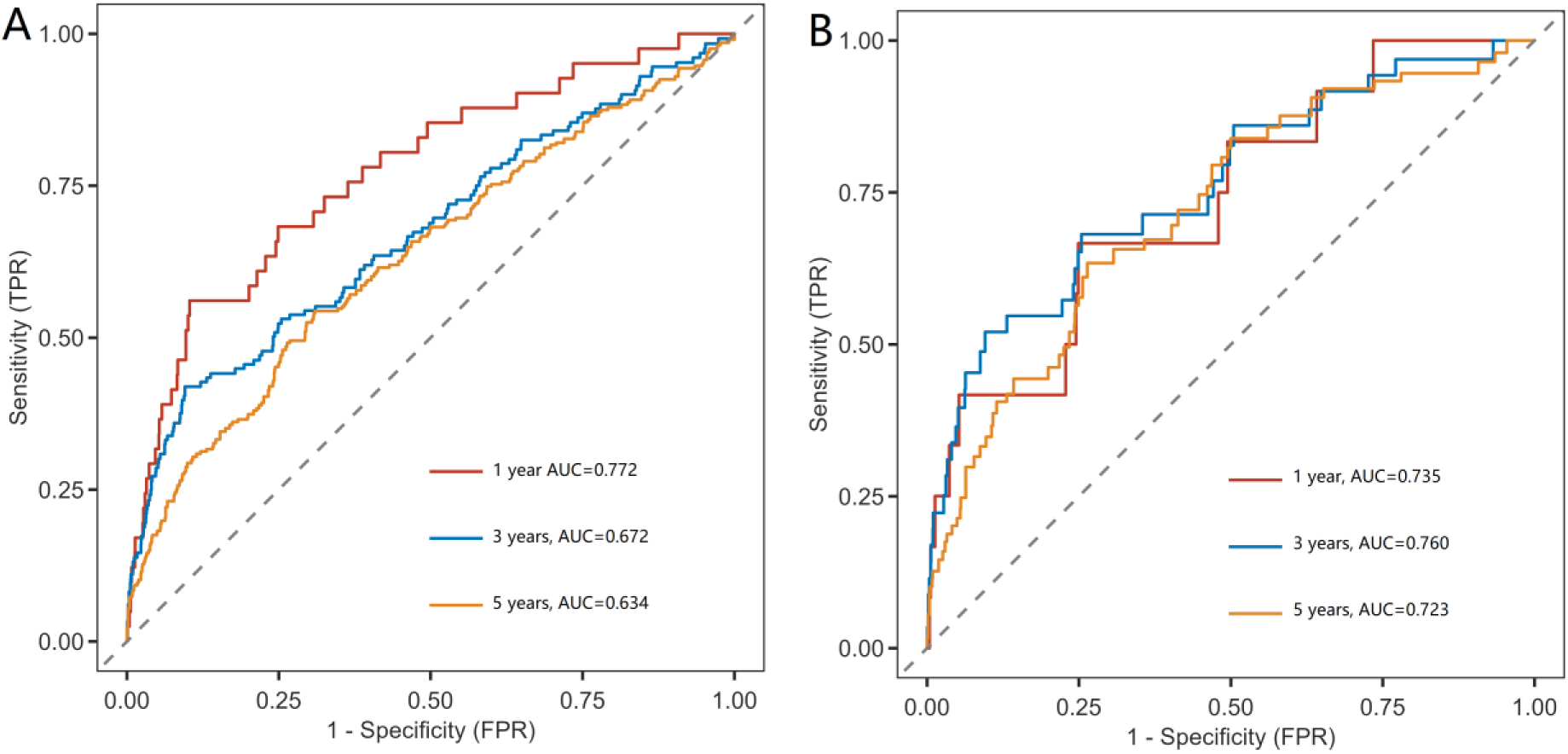
Advanced lung cancer inflammation index (ALI) for predicting overall mortality and cardiovascular mortality among 2805 hypertensive patients, weighted.

### Subgroup analysis

Subgroup analyzes revealed a connection between ALI and both all-cause mortality and cardiovascular mortality, taking into account factors such as age, sex, education level, ethnicity, income, smoking, and diabetes status. No notable interaction was observed between any of these characteristics and ALI in terms of their relationship with all-cause mortality. Interestingly, in terms of the relationship between ALI and cardiovascular mortality, all races in the United States have a better prognosis (Supplemental Material: Figure S4).

## DISCUSSION

The aim of this study was to explore the associations between ALI and all-cause mortality and cardiovascular mortality among hypertensive patients and to predict their risk of death at 1, 3, and 5 years. By analyzing the physical examination data of 2805 hypertensive participants, our research revealed an inverse correlation between ALI and both all-cause mortality and cardiovascular mortality in hypertensive patients and revealed that ALI played a prominent role in predicting their mortality risk. Patients with higher ALI values might experience a lower likelihood of mortality than those with lower ALI values. This relationship was further supported by Kaplan-Meier survival curve analysis, which revealed a clear separation in survival probability between hypertensive patients with high and low ALI levels. Moreover, our study demonstrated that ALI has a robust predictive ability for hypertensive patients at different time points, with area under the curve (AUC) values indicating its effectiveness in predicting both all-cause and cardiovascular mortality at 1, 3, and 5 years. This predictive power was further validated by comparing ALI with the NLR, another inflammation-related biomarker, which showed that ALI was more effective in predicting mortality among hypertensive patients. These findings indicate that ALI can serve as a straightforward and efficient alternative measure for assessing the risk of mortality among hypertensive patients and is convenient for individualized monitoring and management.

To date, only two teams have studied the impact of ALI on the prognosis of hypertensive patients^30,31^. Compared with their research, we divided ALI into four groups from small to large by the equal frequency binning method to achieve more accurate analysis results. We used survey-weighted method to analysis data, which made the analysis results more nationally representative. We found that ALI has a similar effect on reducing patient mortality across different factors through competing risk models. Furthermore, we found the NLR is less effective in predicting mortality than ALI. Previous studies have investigated the relationships between ALI and both all-cause and cardiovascular mortality among various patient groups and the wider population. For example, Chen ^12^ reported that higher ALI levels are linked to lower mortality in type 2 diabetes mellitus (T2DM) patients. Li ^32^ discovered in 2023 that ALI can serve as a novel prognostic biomarker for advanced hepatocellular carcinoma patients undergoing immunotherapy.

Although the specific biological mechanisms by which ALI is associated with mortality among hypertensive patients remain unclear, potential key pathways may involve endothelium impairment and inflammation. As mentioned above “ALI = BMI × Alb/NLR”, and the NLR is calculated from the absolute neutrophil count/absolute lymphocyte count. Neutrophils may promote vascular endothelial injury and the inflammatory response by secreting cytokines such as IL-1β and IL-8, and producing oxidative stress substances such as reactive oxygen species (ROS) and reactive nitrogen species (RNS) ^33,34^. Endothelial impairment or cell death disrupts the vascular barrier, causing vasoconstriction, diastolic dysfunction, proliferation and migration of vascular smooth muscle cells, inflammation, and thrombosis. These effects are tightly linked to the progression of cardiovascular conditions including atherosclerosis, hypertension, coronary artery disease, and cerebral ischemic stroke^35^. Various lymphocyte subsets and their cytokines contribute to vascular remodelling in hypertension, and other heart diseases. Effector T cells, such as Th1 (producing interferon-gamma), Th2 (producing interleukin-4), Th17 (producing interleukin-17), and regulatory T cells (Tregs, expressing Foxp3), exhibit both proinflammatory and anti-inflammatory properties. In particular, Th1 cells may increase blood pressure by directly affecting the kidney and promoting vascular remodelling via cytokines or their impact on perivascular fat. Conversely, adoptive transfer of Tregs can ameliorate oxidative stress in blood vessels and endothelial dysfunction, and reduce the plasma levels of proinflammatory cytokines to prevent increased blood pressure ^36^.

## Strengths and limitations

When considering the results of our study, it is crucial to be aware of both its advantages and disadvantages. Our study was characterized as a large-scale, prospective, population-based study. When investigating the connection between ALI and mortality among patients with hypertension, as well as the ability of ALI to predict mortality, we employed survey-weighted analysis, which makes the research results more nationally representative. Moreover, several limitations should be noted. First, despite adjusting for potential confounding factors, residual confounding factors may still be present. Second, the NHANES did not collect detailed information on hypertension staging, impeding our capacity to perform a more detailed examination of the connection between ALI and mortality across various stages of hypertension. Finally, the participants in this study were all Americans and lacked national representativeness, necessitating further investigations to confirm the applicability of ALI in predicting the prognosis of hypertensive patients globally.

## CONCLUSION

Our study revealed that in the US, ALI has a notable correlation with all-cause mortality and cardiovascular mortality among hypertensive patients, and has strong predictive power for mortality in both the short and long term. Consequently, ALI can be a valuable and effective tool for identifying hypertensive patients who are at heightened risk, and directing tailored interventions. Additional research is needed to validate these conclusions and delve into the fundamental mechanisms involved.

## Data Availability

All data are publicly available and can be accessed at the NHANES (the National Health and Nutrition Examination Survey) website (http://www.cdc.gov/nchs/nhanes/index.htm). Relevant DecisionLinnc and R code is available upon reasonable request to the corresponding author. The first author had full access to all data in the study and takes responsibility for its integrity and the data analysis.

https://www.who.int/data/gho/publications/world-health-statistics.

http://www.cdc.gov/nchs/nhanes/index.htm

## Acknowledgements

The authors thank the NHANES participants and staff for their crucial contributions to this research.

## Source of Funding

This work was supported by a grant from the Health Commission of Hubei Province Funded Project (grant number W2023F043).

## Disclosures

None.

## Supplemental Material

Table S1

Figure S1-S4

## Nonstandard Abbreviations and Acronyms

ALI: Advanced lung cancer inflammation index
NLR: Neutrophil to Lymphocyte ratio
NHANES: National Health and Nutrition Examination Survey

## Notes

### Competing Interest Statement

The authors have declared no competing interest.

### Clinical Trial

All procedures adhered to the principles outlined in the Declaration of Helsinki, and NHANES has secured personal informed and written consent from every participant, ensuring their awareness and agreement in the study.

### Clinical Protocols

http://www.cdc.gov/nchs/nhanes/index.htm

